# Longitudinal Characterization of the Tumoral Microbiome during Radiotherapy in HPV-associated Oropharynx Cancer

**DOI:** 10.1101/2020.06.08.20124974

**Authors:** Houda Bahig, Clifton D Fuller, Aparna Mitra, Travis Solley, Sweet Ping Ng, Ibrahim Abu-Gheida, Baher Elgohari, Andrea Delgado, David I Rosenthal, Adam S Garden, Steven J Frank, Jay P Reddy, Lauren Colbert, Ann Klopp

**Affiliations:** Radiation Oncology Department, University of Texas MD Anderson Cancer Center, Houston, Texas, USA; Radiation Oncology Department, Centre Hospitalier de l’Université de Montréal, Montreal, Quebec, Canada; Radiation Oncology Department, Peter MacCallum Cancer Centre, Melbourne, Australia; Radiation Oncology Department, Burjeel Medical City, Abu-Dhabi, United Arab Emirates; Clinical Oncology and Nuclear Medicine Department, Mansoura University, Mansoura, Egypt

## Abstract

**Purpose:** To describe the baseline and serial tumor microbiome in HPV-associated oropharynx cancer (OPC) over the course of radiotherapy (RT).

**Methods:** Patients with newly diagnosed HPV-associated OPC treated with definitive radiotherapy +/- concurrent chemotherapy were enrolled in this prospective study. Using 16S rRNA gene sequencing, dynamic changes in tumor microbiome during RT were investigated. Surface tumor samples were obtained before RT and at week 1, 3 and 5 of RT. Radiological primary tumor response at mid-treatment was categorized as complete (CR) or partial (PR).

**Results:** Ten patients were enrolled. Mean age was 63 years (range: 51-71). As per AJCC 8^th^ Ed, 50%, 20% and 30% of patients had stage I, II and III, respectively. At 4-weeks, 7 patients had CR and 3 patients had PR; at follow-up imaging post treatment, all patients had CR. Baseline diversity of tumoral and buccal microbiomes was not statistically different. For the entire cohort, alpha diversity was significantly decreased over the course of treatment (p=0.02). There was a significant alteration in the bacterial community within the first week of radiation. Baseline tumor alpha diversity of patients with CR was significantly higher than those with PR (p=0.03). While patients with CR had significant reduction in diversity over the course of radiation (p=0.02), the diversity remained unchanged in patients with PR. Patients with history of smoking had significantly increased abundance of *Granulicatella* (p=0.04), and *Kingella* (0.05) and lower abundance of *Alloprevotella* (p=0.04) compared to never smokers.

**Conclusions:** The tumor microbiome of HPV-associated OPC exhibits reduced alpha diversity and altered taxa abundance over the course of radiotherapy. The baseline bacterial profiles of smokers vs. non-smokers were inherently different. Baseline tumor alpha diversity of patients with CR was higher than patients with PR, suggesting that the microbiome as a biomarker of radiation response deserves further investigation.

## 1. INTRODUCTION

The last decades have seen the emergence of human papillomavirus (HPV) –associated squamous cell carcinoma of the oropharynx cancer (OPC), that now constitutes the vast majority of OPC in North America (1, 2). HPV-associated OPC has a significantly improved prognosis compared to non-HPV-associated OPC (3, 4), and is currently the focus of multiple trials of treatment de-intensification, in an attempt to reduce treatment-related morbidity (5-10). However, it is now recognized that a subset of HPV-associated OPC presents with highly aggressive behaviour and may not be suited for these treatment de-intensification approaches that may result in jeopardized patients outcomes (11). This poor prognosis subgroup remains poorly defined and reliable patient-specific biomarkers that can predict treatment response are greatly needed. Complete response at mid-treatment is observed in as high as 50% of patients with HPV-associated OPC (12), and tumor response to radiotherapy has been shown to be associated with permanent tumor control outcomes (13).

Across multiple cancer sites, cumulating evidence suggests that the microbiome could impact cancer risk and treatment outcomes (14-19). More specifically, the oral microbiome hosts several hundred bacterial species and is emerging as a new potential biomarker reservoir for head and neck cancers (20, 21). Differences in composition between the oral microbiome of patients with oral cavity and oropharynx cancers and that of healthy individuals have lead to recent initiatives investigating the potential role of the oral microbiome for early detection of head and neck malignancies (22, 23). Previous studies have shown that the composition of the oral microbiome was altered over the course of radiotherapy in oral cavity cancer (24, 25). To our knowledge, there is currently no study characterizing the tumor microbiome in OPC treated with radiotherapy.

The aim of this pilot study is two-fold: (1) to describe the baseline and longitudinal changes in tumor microbiome in HPV-associated OPC over the course of radiotherapy (RT); (2) to compare the tumor microbiome of early vs. late radiotherapy responders. In doing so, we hope to generate hypothesis that would lay ground for future larger scale work on the role of tumor microbiome in predicting treatment response.

## 2. MATERIAL AND METHODS

### 2.1 Patients

Patients with newly diagnosed HPV-associated squamous cell carcinoma of the oropharynx treated with radical radiotherapy +/- cisplatin-based chemotherapy and whose tumour could be accessed by surface swab in the clinic were prospectively enrolled. Patients treated with induction chemotherapy or robotic surgery, and patients with previous head and neck malignancy were excluded. Patients were treated with standard of care radiotherapy at a dose of 70 Gy in 33 fractions over 6-7 weeks. This study was approved by our institutional ethics committee (MDACC 2014-0543), and all patients signed a consent form.

### 2.2 Response Assessment

All patients underwent pre-treatment planning computed tomography (CT) and, unless contra-indicated, a planning magnetic resonance imaging (MRI) of the head and neck region. In addition, all patients underwent mid-treatment (week 4) repeat planning CT and MRI for early primary tumor response assessment. Mid-treatment primary tumor response was classified as complete (CR), partial (PR) or stable (SD), based on RECIST criteria version 1.1.. A contrast-enhanced CT of the head and neck was obtained at 8 weeks after treatment completion for definitive treatment response assessment. In cases where response at 8 weeks post treatment was incomplete, a fluorodeoxyglucose (FDG)-positron emission tomography/ computed tomography (PET/CT) was obtained at 12 weeks after RT completion for further assessment of response.

### 2.3 Microbiome sample collection and analysis

Tumor microbiome samples were obtained immediately before RT start and at week 1, 3 and 5 using a matrix designed quick release Isohelix swab (DSK-50 and XME-50, isohelix.com, UK). Samples were obtained by brushing the swab against the viable tumor several times in order to collect enough cells from the tumor surface. At baseline (i.e. immediately before RT start), an oral microbiome swab was also obtained by brushing the swab against healthy buccal mucosa, to serve as control. Isohelix swabs were placed in 20□μL of protease K and 400□μL of lysis buffer (Isohelix) and stored at −80□°C within 1□h of sample collection. Bacterial genomic DNA was extracted using MO BIO PowerSoil DNA Isolation Kit (MO BIO Laboratories). Microorganism identification derived from culture-based approaches, using the 16 Svedberg unit (S) rRNA gene technology for bacteria. 16S rRNA gene sequencing was performed by the Alkek Center for Metagenomics and Microbiome Research at Baylor College of Medicine. 16S rRNA was sequenced methods previously detailed in the Human Microbiome Project (26). Shannon diversity index was used to evaluate alpha (within sample) diversity. Relative abundance of the microbiome taxa and genera was compared between patients presenting CR vs. PR/SD at mid-treatment. Using the Kruskal-Wallis test, an α of ≤0.05 was considered significant.

## 3. RESULTS

### 3.1 Patients characteristics

Ten patients were enrolled. Mean age was 63 years (range: 51-71) and all patients were male **(Table 1)**. Tumor was located in the tonsil and in the base of tongue in 60% and 40% of patients, respectively. As per AJCC 8^th^ Ed, 50%, 20% and 30% of patients had stage I, II and III, respectively. 40% were smokers (active or past), while 60% never smoked. All patients were treated to a dose of 70 Gy in 33-35 fractions. Eight patients underwent concurrent platin-based chemotherapy that consisted in weekly cisplatin at 40 mg/m^2^, and one patient received Nivolumab every 14 days for 10 doses starting 14 days prior to IMRT. At 4-weeks, 7 patients had CR and 3 patients had PR. At 8 weeks post treatment, all patients with CR at mid treatment had persistent CR, while among patients with mid-treatment PR, 1 had CR and 2 had persistent PR at the primary site and/or lymph nodes. All patients had complete response on PET/CT at 12 weeks post-treatment. Six patients were treated with volumetric arc therapy while 4 patients were treated with intensity modulated proton therapy.

**Table 1.**
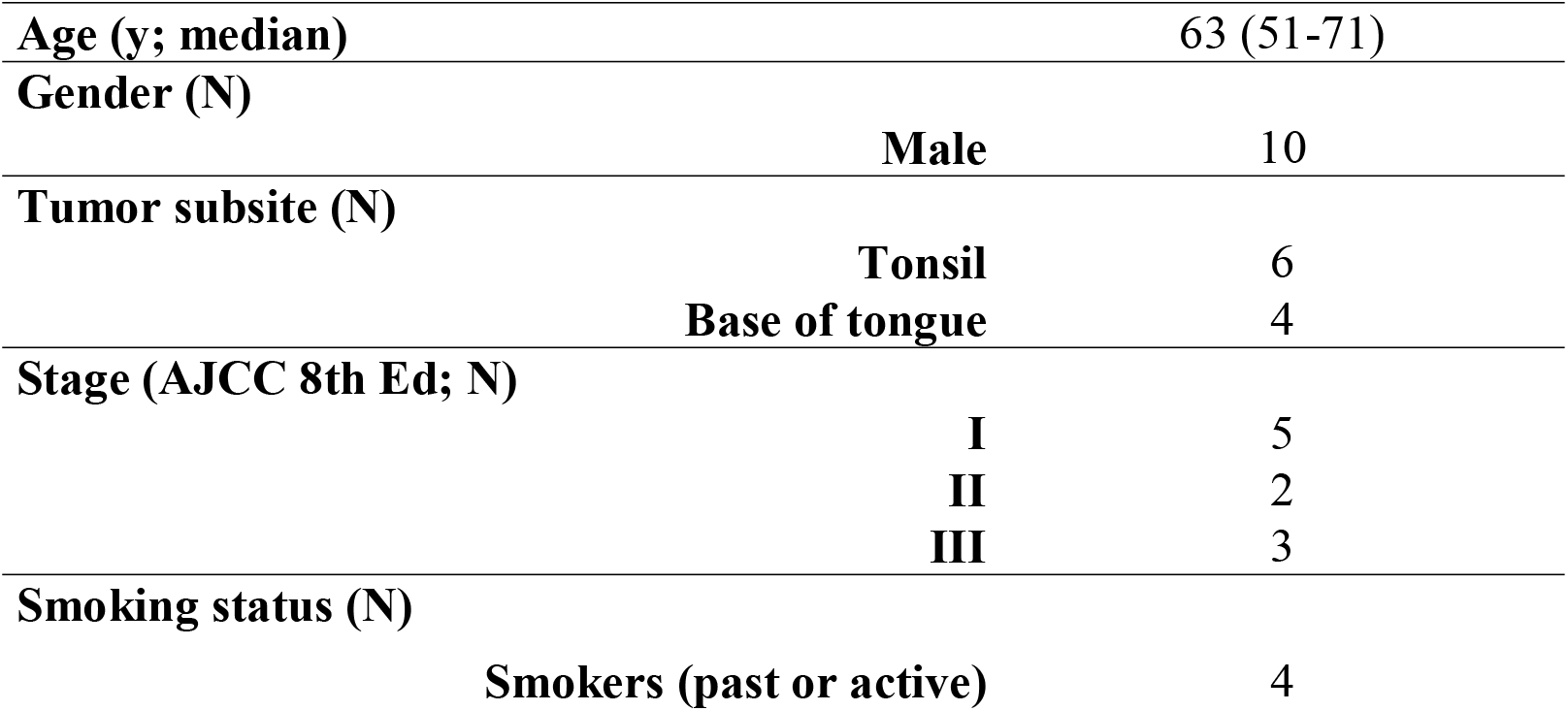
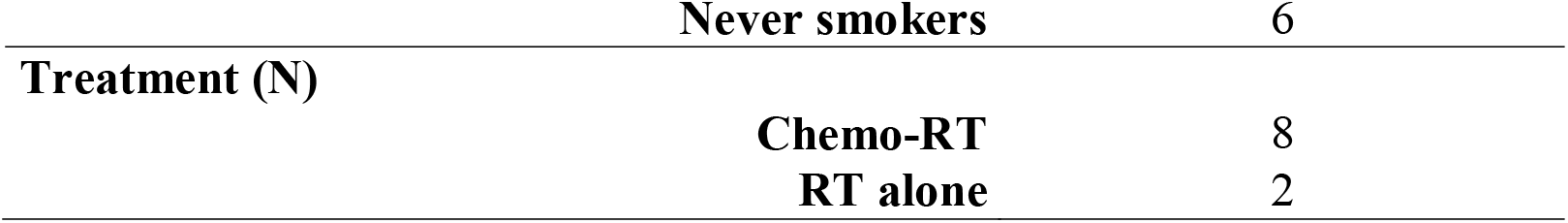
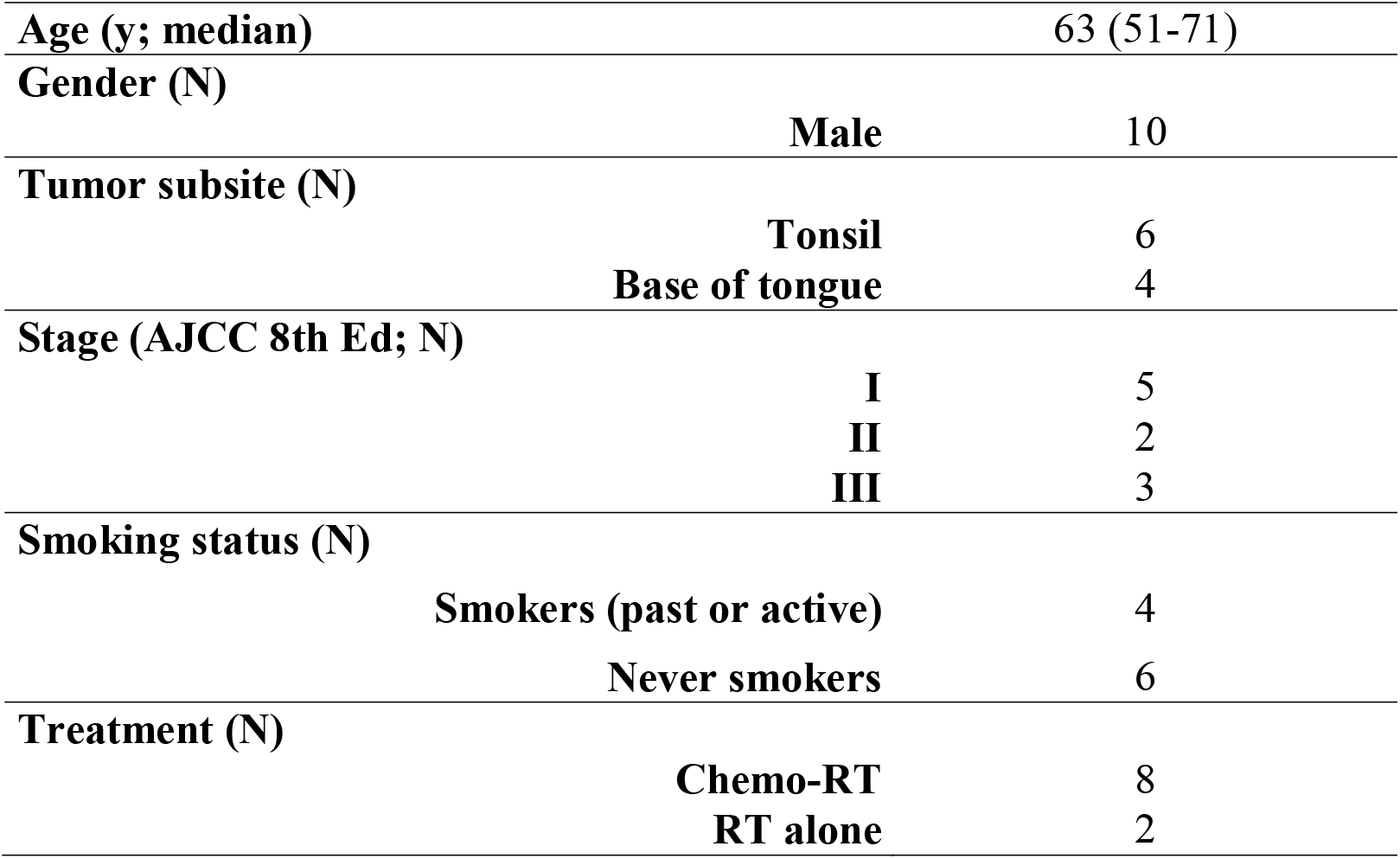
Patients Characteristics.

### 3.2 Characterisation of the mucosal microbiome

Baseline alpha diversity of tumoral and buccal microbiomes was not statistically different (**Figure 1**). Baseline alpha diversity of patients who had a CR by 4 weeks was significantly higher than those who had a PR (p=0.03) (**Figure 2**). For the entire cohort, alpha diversity was significantly decreased over the course of treatment (p=0.02) (**Figure 3**). However, patients with CR at 4-weeks had significantly reduced alpha diversity over the course of radiation (p=0.02), while diversity remained unchanged in patients with PR. Patients with history of smoking had significantly increased abundance of *Granulicatella* (p=0.04), and *Kingella* (0.05) and lower abundance of *Alloprevotella* (p=0.04) compared to never smokers **(Figure 4)**. There was a shift in the composition of the bacterial community over the course of radiation. The relative composition of Actinomyces declined over the course of radiation. Other organisms, such as *Gemella* and *Streptococcus*, decreased in abundance between baseline and week 1 and subsequently returned to baseline abundance by week 5. **(Figure 5)**. This was followed by progressive re-normalization on subsequent weeks.

**Figure 1.**
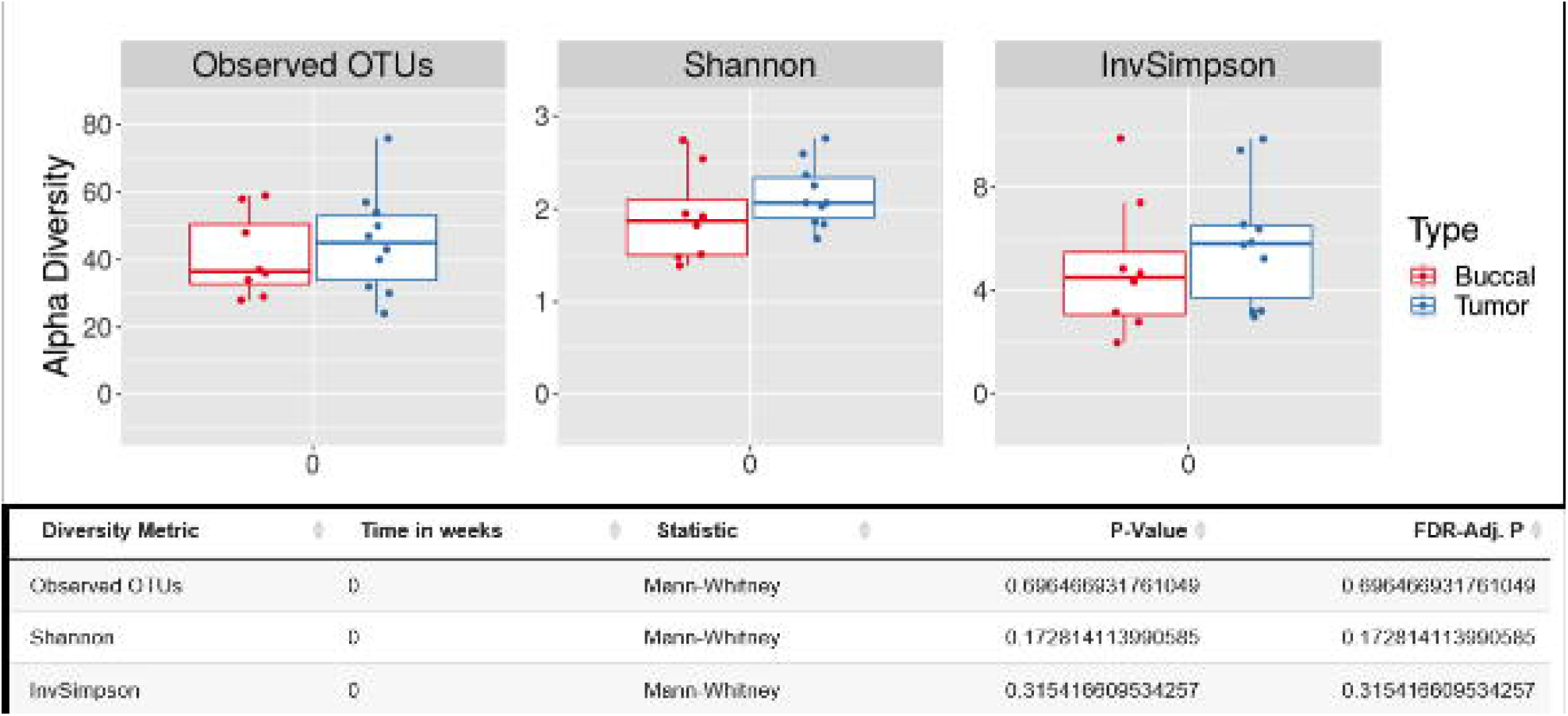
Baseline alpha diversity of tumoral and buccal microbiomes.

**Figure 2.**
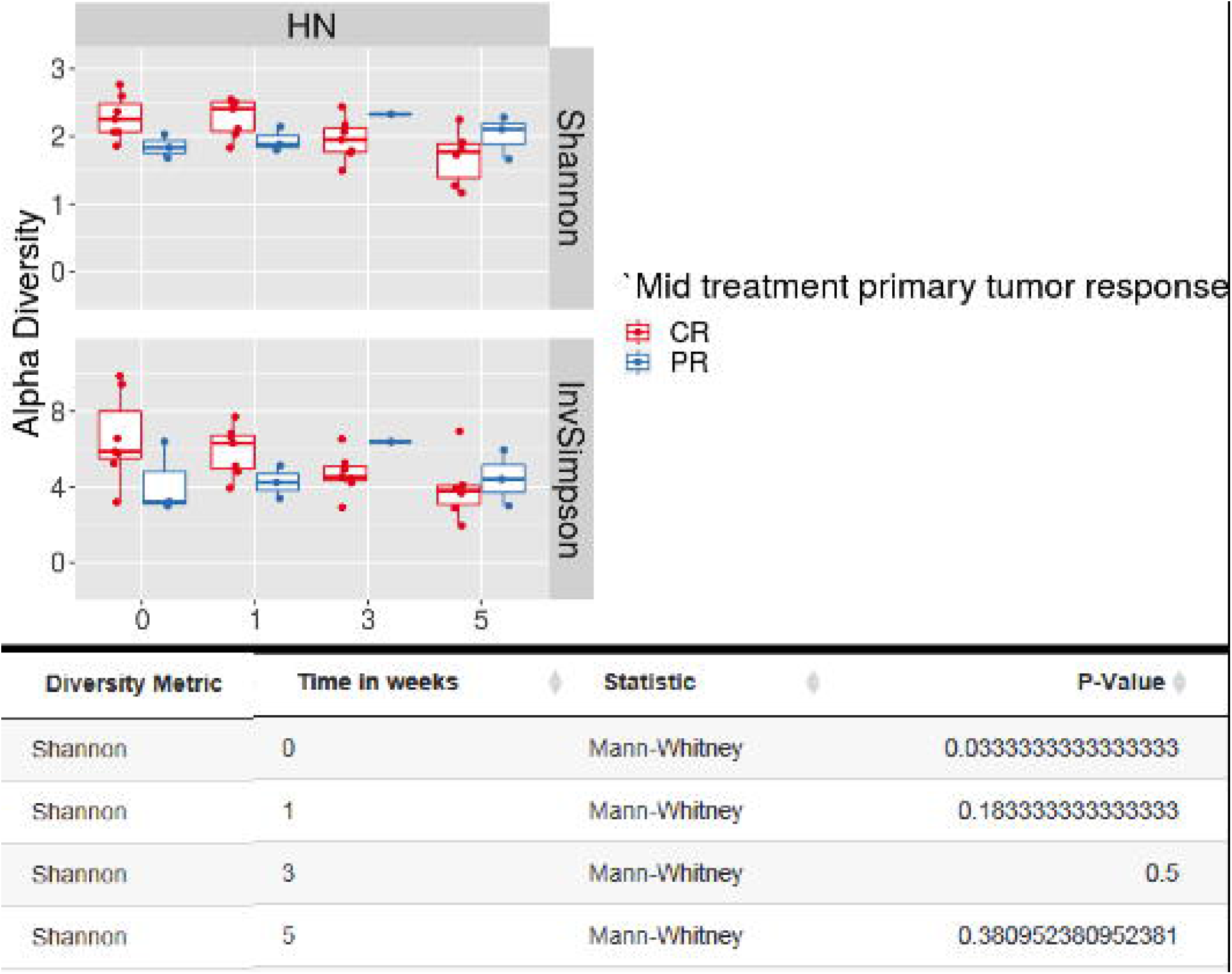
Baseline alpha diversity of patients with CR vs. PR at mid-treatment.

**Figure 3.**
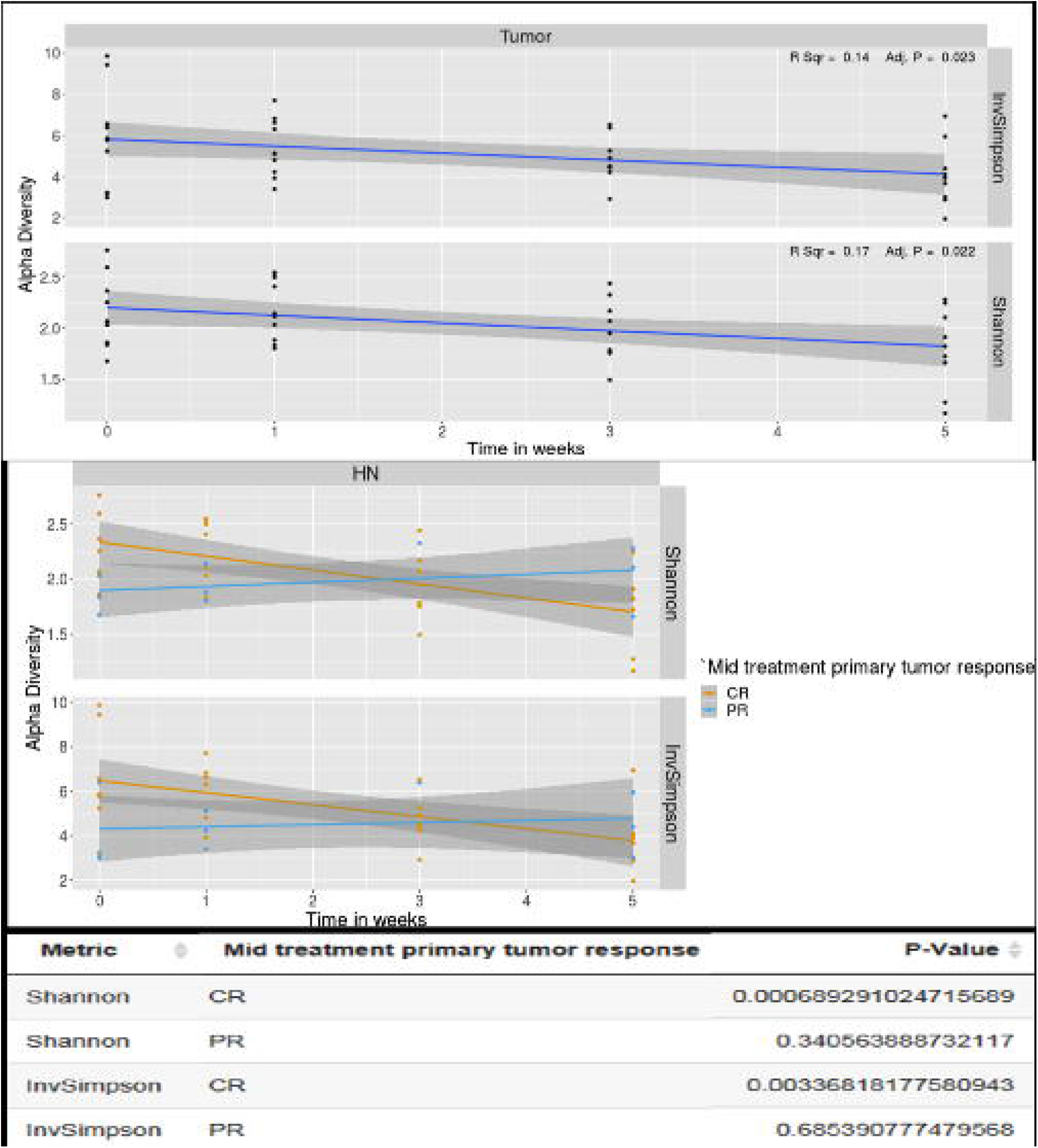
Changes in alpha diversity over the course of treatment.

**Figure 4.**
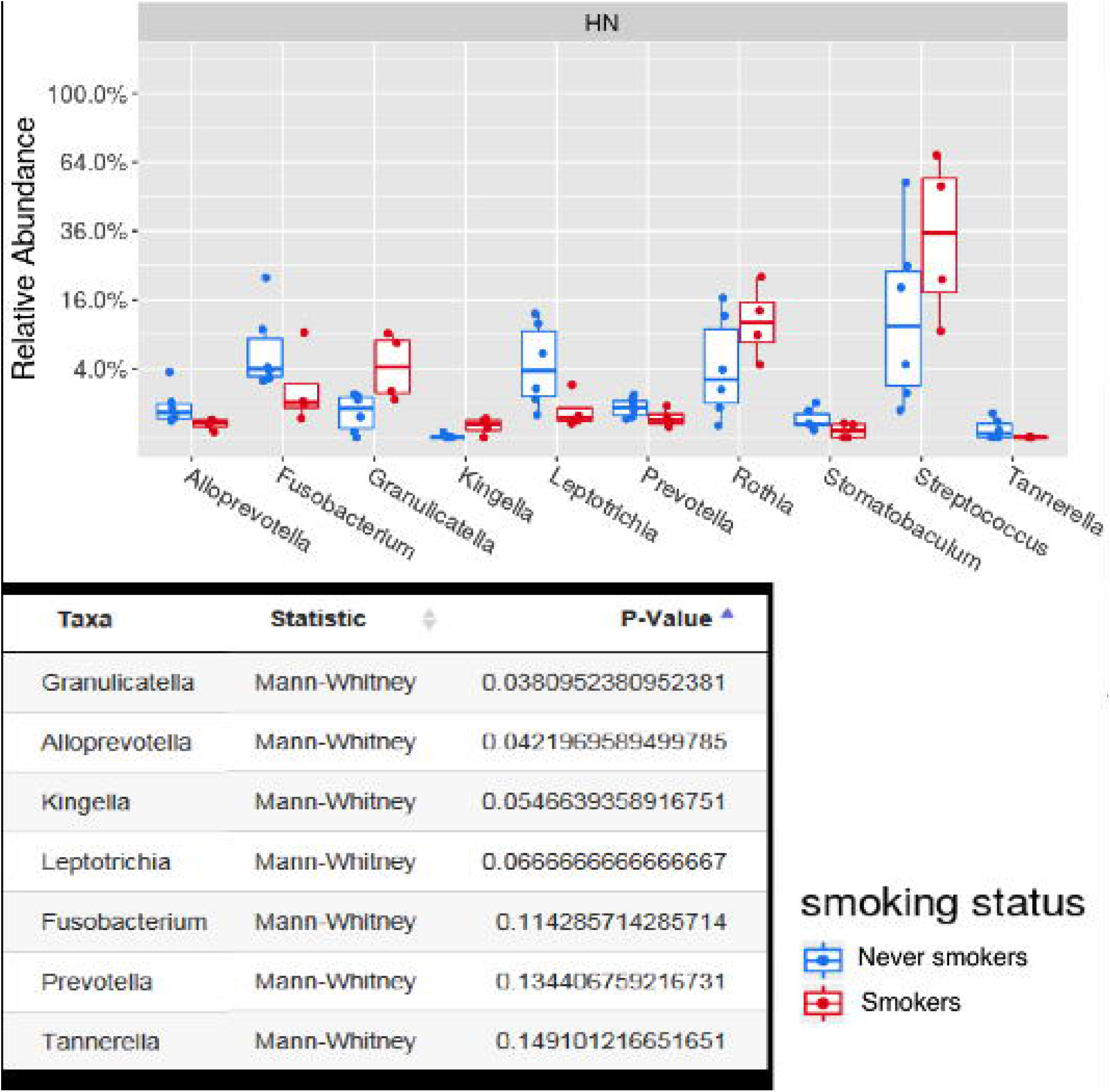
Comparison of the microbial composition of smokers vs. never smokers.

**Figure 5.**
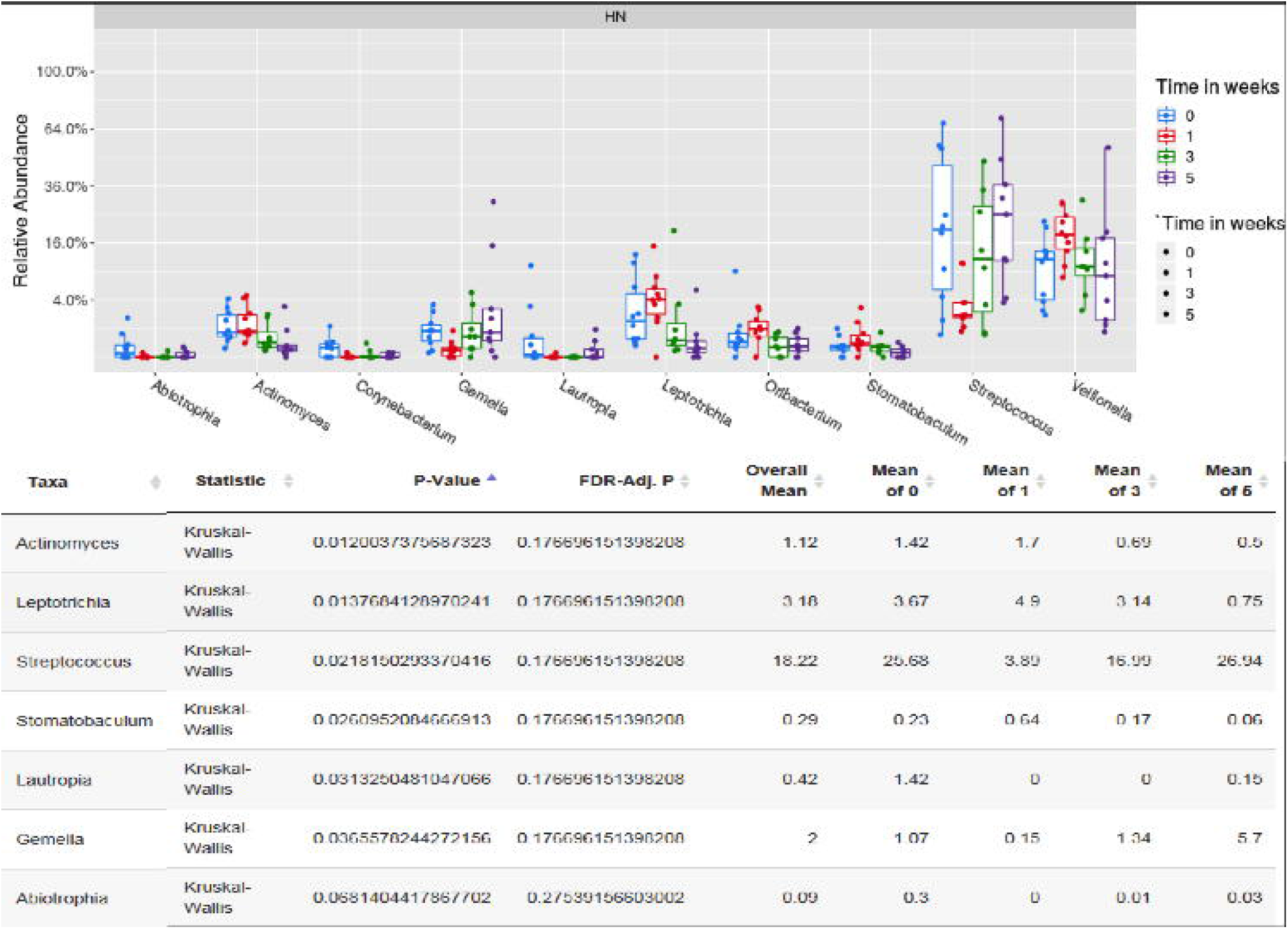
Dynamic alteration in taxa abundance over the course of radiation. The relative abudance of organisms at the genus level at baseline and throughout the course of chemoradiation are shown.

## 4. DISCUSSION

This is the first study characterizing the tumor microbiome in HPV-associated OPC and exploring its role in the prediction of response to radiotherapy. In this pilot study, we report that the alpha diversity was significantly reduced over the course of treatment and that the composition of the microbiota was rapidly altered as early as in the first week of radiation. The baseline alpha diversity of the tumor microbiome was significantly higher in rapid responders (i.e. those with complete response at mid-treatment). We found that the tumor microbiome profiles of smokers vs. non-smokers were significantly different.

The oral microbiome has been hypothesised to play a role in the pathogenesis of head and neck cancers, through chronic inflammation, anti-apoptotic cellular signalling and release of carcinogens (27). In fact, inherent differences in the bacterial flora of healthy individuals versus individuals with oral cavity/oropharynx cancers have been documented in several studies, and various mechanisms supporting a carcinogenic role of the oral microbiome have been postulated (28-33). As an example, acetaldehyde, a toxic ethanol metabolite produced via bacterial oxidation is found in higher concentration in heavy alcohol users with poor oral hygiene and its accumulation has been linked to increased risk of oral cavity cancer (34, 35). Similarly, a synergistic effect of periodontitis - a chronic infection caused by gram-negative anaerobic bacteria- and oral HPV co-infection has been suggested for the development of OPC cancer, as the continuous release of bacterial toxins and inflammatory markers in the oral cavity would enhance epithelial tissue abrasion, basal cells infection, and ultimately persistence of HPV infection (36). The role of the oral microbiome to predict the therapeutic efficacy of radiotherapy in patients with head and neck cancers has never previously been investigated. Nonetheless, consistent with the results of the present study, our group has previously shown that an increased diversity of the vaginal microbiome was also associated with enhanced response to radiotherapy in HPV-associated cervical cancer (37). In addition, numerous recent studies have focused on the role the gut microbiome as a modulator of the response to immune checkpoint inhibitors in several cancers, including melanoma, lung and renal cancer (14-19). In these studies, an increased alpha diversity of the microbiome was associated with higher therapeutic efficacy of immune checkpoint inhibitors (14, 19). However, in Checkmate 141, a trial investigating the role of Nivolumab in recurrent or metastatic head and neck cancer, the oral microbiome and its alpha diversity were not predictive of the therapeutic response to immunotherapy (38). A study at the Princess Margaret Cancer Centre (NCT034106615) assessing oral and intestinal microbiota in patients with head and neck cancer treated with radical chemoradiotherapy is currently on-going.

The dynamic variation of the oral microbiome over the course of head and neck radiotherapy has not been well studied. Huo et al, correlated the dynamic oral microbiome change with the severity of mucositis in patients treated for nasopharynx cancer and found that, although the bacterial alpha diversity did not vary significantly over treatment, there were synchronous shifts in the abundance of specific bacteria such as Prevotella, Fusobacterium, Treponema and Porphyromonas (25). Similarly, Zhu et al. showed that the microbiome profile of patients undergoing radiotherapy for nasopharynx cancer was progressively altered during radiation therapy, in close correlation with development of mucositis (24, 25).

Our study reports preliminary outcomes from an on-going initiative assessing the role of the microbiome in HPV-associated cancers. Given the small sample size, the results of this report, while hypothesis generating, are not definitive. Further work is required to further understand and determine if the tumor microbiome truly has a role in the prediction of therapeutic efficacy in head and neck cancers. In addition, distinctions between the dynamic alterations of the microbiome of patients treated with radiotherapy alone vs. concurrent chemoradiation could not be effectively studied. Another limitation is the lack of concurrent gut microbiome data; however, the role of the gut microbiome may not have the same importance in the context of head and neck radiotherapy as compared to systemic therapy, given that the main anti-tumor mechanisms are local. Finally, an inherent limitation of studies assessing the role of the microbiome is the regional microbiome variations, which may limit the external validity of study findings and warrant local confirmatory investigations.

In conclusion, we found that the tumor microbiome of our entire cohort of patients with HPV-associated OPC exhibits reduced alpha diversity over the course of radiation. This was also associated with an altered taxa abundance manifesting as early as in the first week of radiotherapy. Although not definitive, we found that baseline alpha diversity of patients with complete response by mid-treatment was higher than patients with partial responses or stable disease. While the diversity of the tumor microbiome as a biomarker of radiation response in OPC deserves further investigation, these preliminary outcomes are encouraging and lay ground for further work in that direction.

## Data Availability

Data available upon request

